# Eleven key measures for monitoring general practice clinical activity during COVID-19 using federated analytics on 48 million adults’ primary care records through OpenSAFELY

**DOI:** 10.1101/2022.10.17.22281058

**Authors:** Louis Fisher, Helen J. Curtis, Richard Croker, Milan Wiedemann, Victoria Speed, Christopher Wood, Andrew Brown, Lisa EM Hopcroft, Rose Higgins, Jon Massey, Peter Inglesby, Caroline E. Morton, Alex J. Walker, Jessica Morley, Amir Mehrkar, Seb Bacon, George Hickman, Orla Macdonald, Tom Lewis, Marion Wood, Martin Myers, Miriam Samuel, Robin Conibere, Wasim Baqir, Harpreet Sood, Charles Drury, Kiren Collison, Chris Bates, David Evans, Iain Dillingham, Tom Ward, Simon Davy, Rebecca M. Smith, William Hulme, Amelia Green, John Parry, Frank Hester, Sam Harper, Jonathan Cockburn, Shaun O’Hanlon, Alex Eavis, Richard Jarvis, Dima Avramov, Paul Griffiths, Aaron Fowles, Nasreen Parkes, Brian MacKenna, Ben Goldacre

## Abstract

**Background:** The COVID-19 pandemic has had a significant impact on delivery of NHS care. We have developed the OpenSAFELY Service Restoration Observatory (SRO) to describe this impact on primary care activity and monitor its recovery.

**Objectives:** To develop key measures of primary care activity and describe the trends in these measures throughout the COVID-19 pandemic.

**Methods:** With the approval of NHS England we developed an open source software framework for data management and analysis to describe trends and variation in clinical activity across primary care electronic health record (EHR) data on 48 million adults.

We developed SNOMED-CT codelists for key measures of primary care clinical activity selected by a expert clinical advisory group and conducted a population cohort-based study to describe trends and variation in these measures January 2019-December 2021, and pragmatically classified their level of recovery one year into the pandemic using the percentage change in the median practice level rate.

**Results:** We produced 11 measures reflective of clinical activity in general practice. A substantial drop in activity was observed in all measures at the outset of the COVID-19 pandemic. By April 2021, the median rate had recovered to within 15% of the median rate in April 2019 in six measures. The remaining measures showed a sustained drop, ranging from a 18.5% reduction in medication reviews to a 42.0% reduction in blood pressure monitoring. Three measures continued to show a sustained drop by December 2021.

**Conclusions:** The COVID-19 pandemic was associated with a substantial change in primary care activity across the measures we developed, with recovery in most measures. We delivered an open source software framework to describe trends and variation in clinical activity across an unprecedented scale of primary care data. We will continue to expand the set of key measures to be routinely monitored using our publicly available NHS OpenSAFELY SRO dashboards with near real-time data.

## Background

The COVID-19 pandemic has significantly affected the capacity and delivery of both primary and secondary care within the NHS ^1–4^. We have previously described a data-driven approach to analyse, review and prioritise activity in NHS primary care in collaboration with a clinical advisory group through the establishment of the OpenSAFELY *NHS Service Restoration Observatory* (SRO) ^5,6^. Following the first wave in March 2020, we found that some clinical activities were not restored to near normal levels by December 2020 as was anticipated in guidance issued by NHS England in July 2020^7^. This entailed a vast volume of data analysis, likely in excess of what could be realistically monitored by clinical and commissioning teams. Informed by this work and in collaboration with our clinical advisory group we suggested key measures of primary care clinical activity to support routine monitoring, targeted action and inform response to the COVID-19 pandemic^5,6^.

OpenSAFELY is a secure analytics platform for electronic patient records built by our group on behalf of NHS England to deliver urgent academic and operational research during the pandemic. Our prior work analysed clinical codes recorded across all primary care records from the Electronic Health Record (EHR) vendor TPP, covering 40% of all general practices in England. We have since extended the OpenSAFELY platform to both major EHR vendors, TPP and EMIS, allowing federated analyses and dashboards to be executed across the full primary care records for all patients registered at 99% of England’s practices.

We therefore set out to: develop the proposed key measures of activity from our previous work; describe trends and variation in these measures; assess recovery of service after the first year of the pandemic; and extend the analysis to cover the 48 million adults’ records available using OpenSAFELY.

## Methods

### Study design

We conducted a retrospective cohort study using GP primary care EHR data from all England GP practices supplied by the EHR vendors TPP and EMIS.

### Data sharing

All data were linked, stored and analysed securely within the OpenSAFELY platform: https://opensafely.org/. Data include pseudonymised data such as coded diagnoses, medications and physiological parameters. No free text data are included. All code is shared openly for review and re-use under MIT open license (https://github.com/opensafely/SRO-Measures). Detailed pseudonymised patient data is potentially re-identifiable and therefore not shared.

### Study population

We included all adult patients (n=48,352,770) who were alive and registered with a TPP or EMIS general practice (n=6,389 practices) in England at the beginning of each month between January 2019 and December 2021. All coded events in each month for each monthly cohort were included. We also identified demographic variables for these patients including age, sex, region of their practice address, index of multiple deprivation (IMD) and ethnicity.

### Key measures of clinical activity

#### Development of key measures

In order to develop key measures of NHS clinical activity we convened a clinical advisory group made up of: front-line general practitioner and pharmacists; national clinical advisors and pathology leads; and clinical and research staff from the Bennett Institute. This group manually reviewed charts representing coding of clinical activity the development of which we have described in detail elsewhere ^5,6^. Briefly, we used the CTV3 terminology coding hierarchy (the coding system available in OpenSAFELY-TPP at the time) to produce a large number of charts indicating variation in clinical coding activity between practices across a range of clinical areas. For each clinical area, these charts were manually reviewed by the clinical advisory group in a series of online meetings to prioritise clinical topics that would benefit from routine monitoring and targeted action. The clinical advisory group was asked to suggest key measures for each clinical area considering the following criteria: high volume usage, clinically relevant to front-line practice and whether they are more widely indicative of other problems in service delivery across the NHS (for example a decrease in records for blood tests for kidney function may be a true drop in GPs requesting these tests or it may be related to delays in laboratories processing the results).

The Bennett Institute team took these suggested measures and manually curated bespoke lists of codes (see below). Charts of the newly developed measures were then presented back to the clinical advisory group for a final review alongside a “why it matters” text (Table 1), indicating why each measure is important to monitor.

**Table 1.**
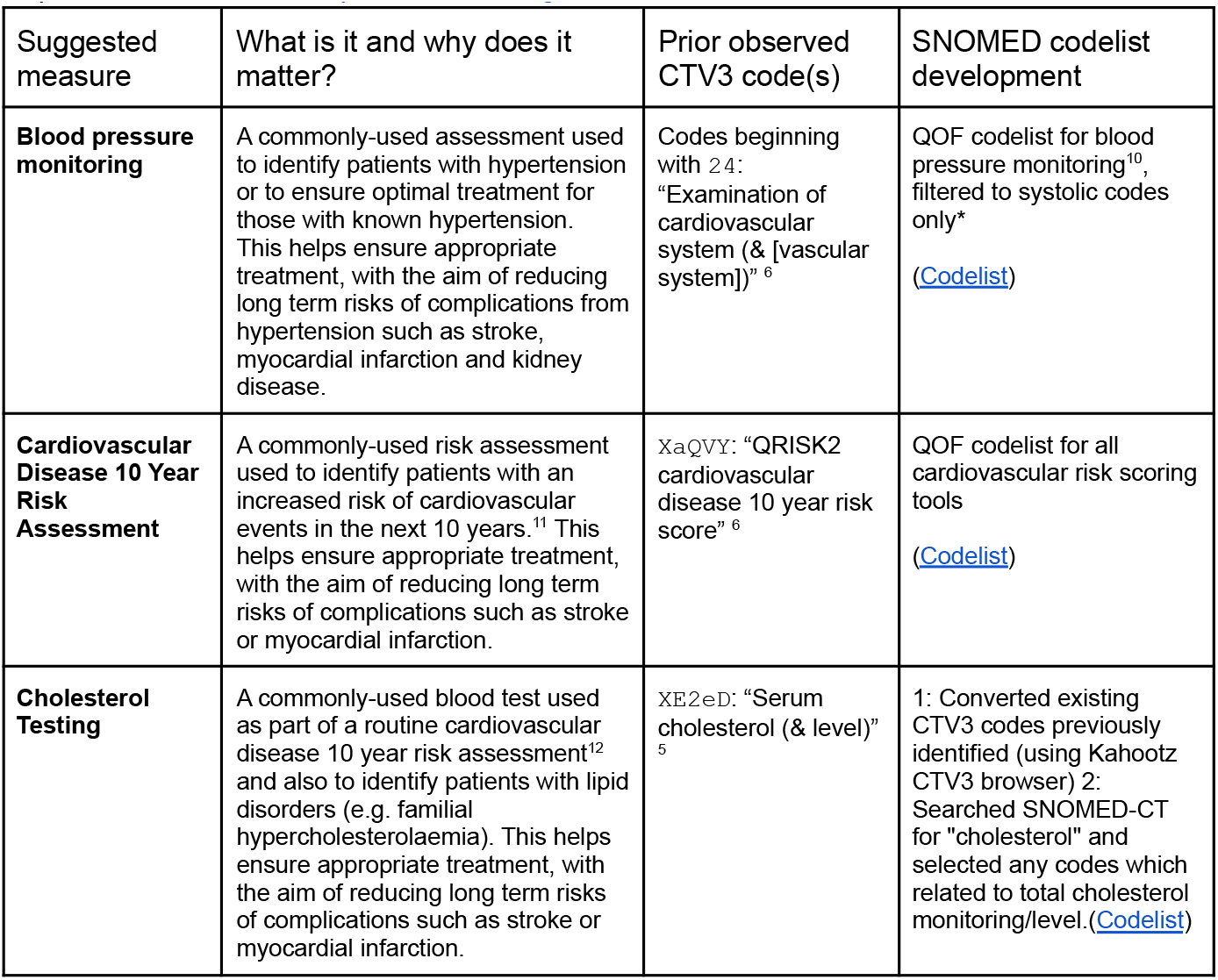
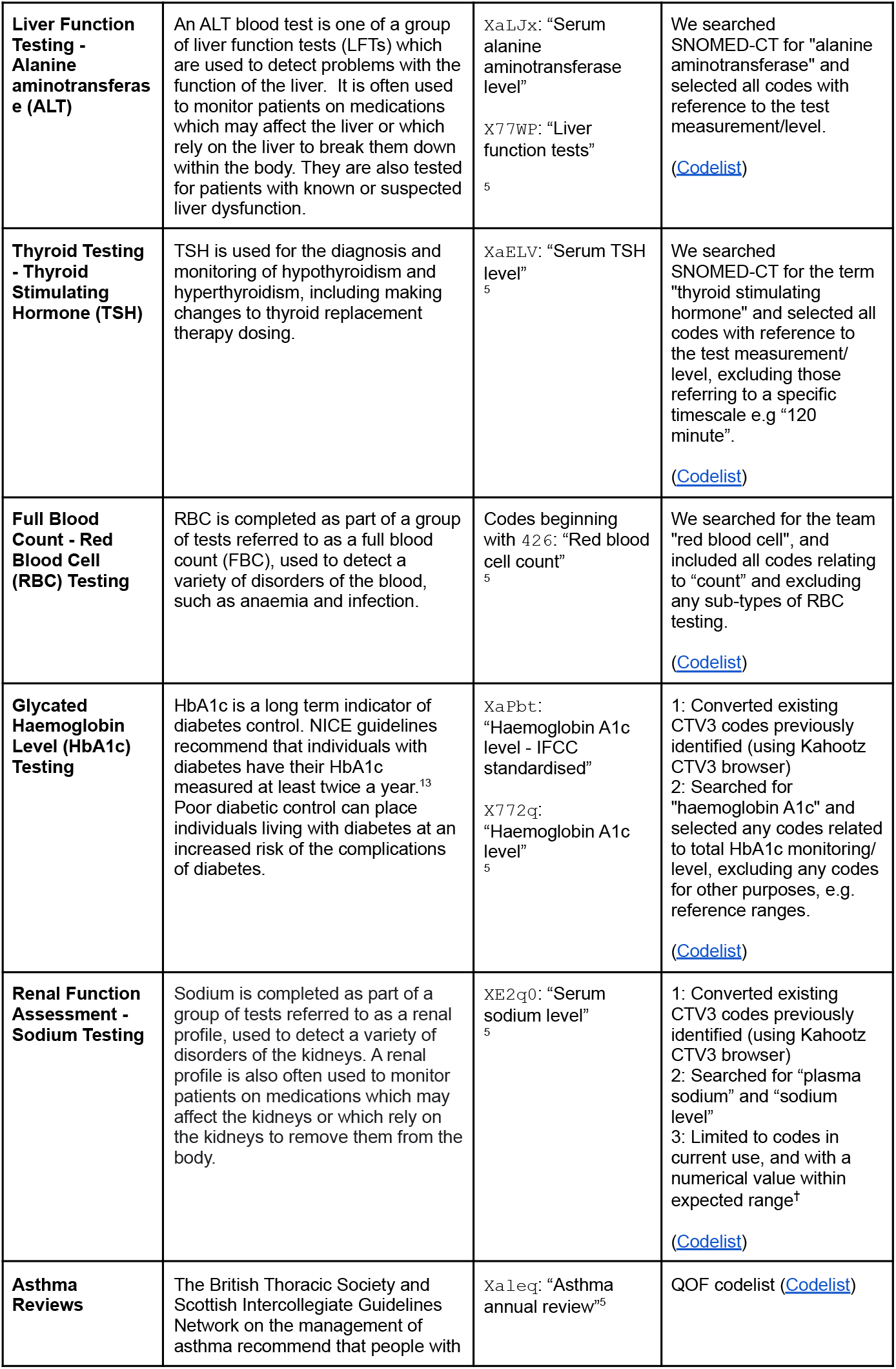
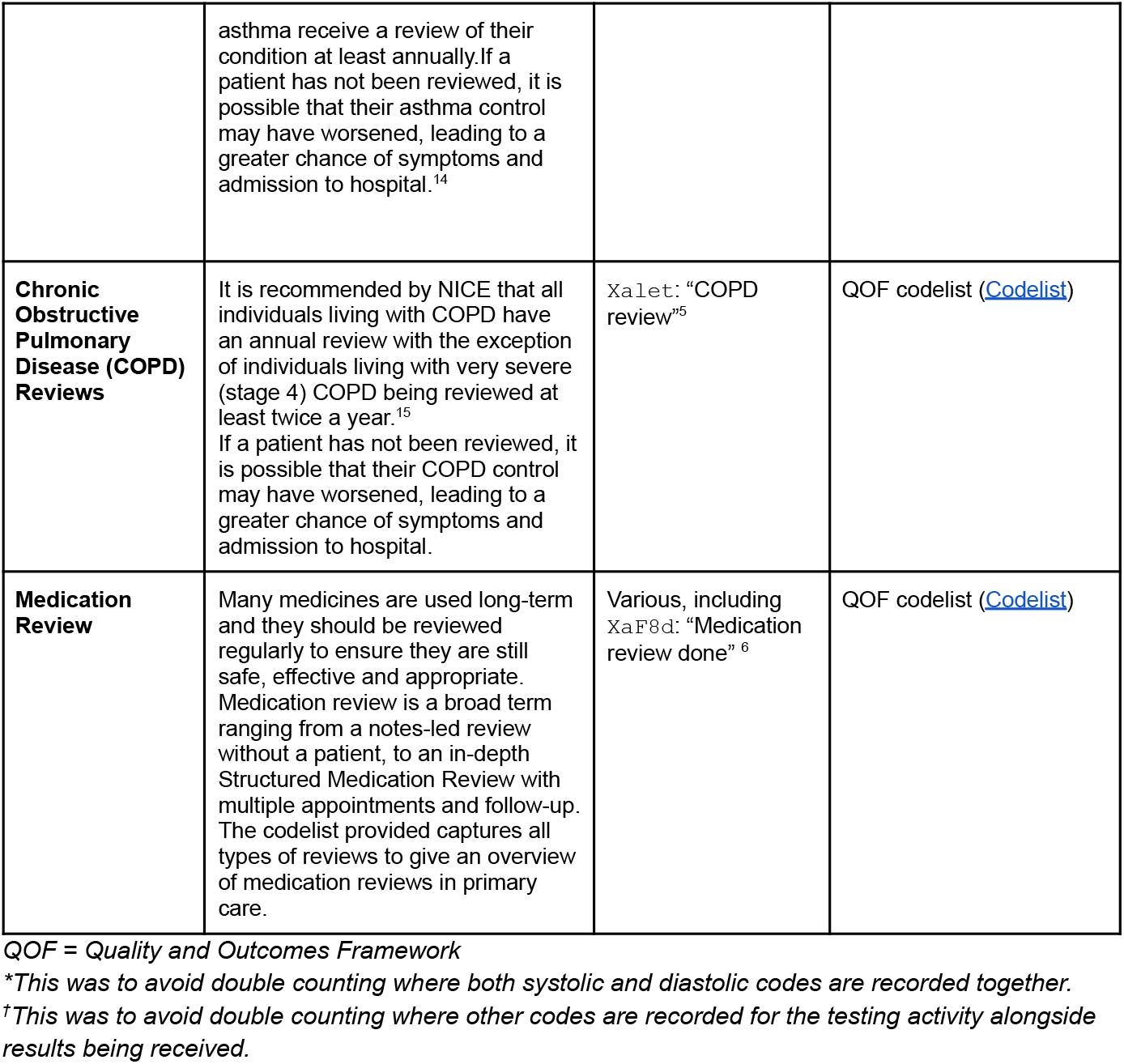
Development of key measures and their associated codelists. A link to each codelist used to define the final key measure is given; all codelists are openly available for inspection and re-use at opencodelists.org.

This measure development process was a pragmatic one based on our experience developing measures for the OpenPrescribing platform^8^, an online viewer of GP prescribing patterns with 20,000 unique users and 100 measures of clinical effectiveness, cost effectiveness and safety. These measures have been iterated over time according to feedback around clinical utility and changes in service delivery. Similarly, we anticipate that we will continue to develop and expand the measures developed here.

#### Codelists

For each key measure of activity, we used the SNOMED-CT coding system, as the mandated NHS standard, to develop a single codelist (Table 1), which can be deployed across any system using SNOMED-CT. Where a well-defined nationally curated codelist existed, such as Quality and Outcomes Framework (QOF), we used the codelist released by NHS Digital^9^. For pathology testing measures with no NHS mandated codelists, we searched for SNOMED-CT codes using OpenCodelists. For this proof of concept we pragmatically decided not to implement all additional complex logic and exceptions that may be associated with national schemes.

#### Data Processing

For each measure we calculated the monthly rate of coding activity per 1000 registered adults for each practice. Where multiple codes from a single codelist are recorded in the patient record in a single month only a single record will be returned. This is advantageous where a practice may use multiple codes to document a single broad activity carried out on a single day, for example recording *Asthma annual review* (SNOMED code 394700004) and a component of the review *Asthma medication review* (394720003) but will not capture two genuine activities carried out at a different time in the same month, for example blood tests measured three weeks apart.

We excluded practices not recording a single instance of a relevant code in each codelist across the entire study period from further analysis;as all the measures analysed here are high volume, any practices with zero recorded events for a measure are likely atypical. We counted the number of practices using each codelist, as well as the total number of unique patients with events across the entire study period, and the total number of events they each experienced (with each patient contributing a maximum of 1 event per month). We then calculated the median and deciles of coding activity rates across all practices each month.

We present the data in time trend decile charts, which we make openly available at reports.opensafely.org and can update regularly.

#### Classification of service restoration

For each key measure chart we classified the change in coding activity using the median rate in April 2020 and 2021 compared to April 2019 which we defined as the “baseline”. April was identified as the first full month of a full “lockdown” in England in 2020 and additionally had 20 business days in 2019, 2020 and 2021, allowing fair comparison. The classification system (Box 1), extends previously developed methods to classify change from baseline based on percentage changes^5^.

##### Box 1

Service change classification relative to baseline (April 2019)

- For April 2020 and April 2021
  - **no chang**e: activity remained within 15% of the baseline level;
  - **increase**: an increase of >15% from baseline;
  - **decrease**: a decrease of >15% from baseline;
- Overall classification:
  - **no change**: no change in both April 2020 and April 2021;
  - **increase**: an increase in April 2020 and April 2021;
  - **temporary increase**: an increase in April 2020 which returned to no change by April 2021.
  - **delayed increase**: no change in April 2020 and an increase in April 2021.
  - **delayed decrease**: no change in April 2020 and a decrease in April 2021.
  - **sustained drop**: a decrease in April 2020 which did not return to no change by April 2021;
  - **recovery**: a decrease in April 2020, which returned to no change by April 2021.

### Software and Reproducibility

Data management and analysis was performed using the OpenSAFELY software libraries and Python, both implemented using Python 3.8 with all code shared openly for review and reuse github.com/opensafely/SRO-Measures. All codelists used are openly available for inspection and re-use at OpenSAFELY Codelists ^16^. This analysis was delivered using federated analysis through the OpenSAFELY platform: codelists and code for data management and data analysis were specified once using the OpenSAFELY tools; then transmitted securely to the OpenSAFELY-TPP platform within TPP’s secure environment, and separately to the OpenSAFELY-EMIS platform within EMIS’s secure environment, where they were each executed separately against local patient data; summary results were then reviewed for disclosiveness, released, and combined for the final outputs. All code for the OpenSAFELY platform for data management, analysis and secure code execution is shared for review and re-use under open licenses at github.com/opensafely-core.

### Patient and Public Involvement

This analysis relies on the use of large volumes of patient data. Ensuring patient, professional, and public trust is therefore of critical importance. Maintaining trust requires being transparent about the way OpenSAFELY works, and ensuring patient and public voices are represented in the design and use of the platform. Between February and July 2022 we ran a six month pilot of Patient and Public Involvement and Engagement activity designed to be aligned with the principles set out in the Consensus Statement on Public Involvement and Engagement with Data-Intensive Health Research ^17^. Our engagement focused on the broader OpenSAFELY platform and comprised three sets of activities: explain and engage, involve and iterate and participate and promote. To engage and explain, we have developed a public website at opensafely.org that provides a detailed description of the OpenSAFELY platform in language suitable for a lay audience and are co-developing an accompanying explainer video. To involve and iterate we have created the OpenSAFELY ‘Digital Critical Friends’ Group; comprised of approximately 12 members representative in terms of ethnicity, gender, and educational background, this group has met every 2 weeks to engage with and review the OpenSAFELY website, governance process, principles for researchers and FAQs. To participate and promote, we are conducting a systematic review of the key enablers of public trust in data-intensive research and have participated in the stakeholder group overseeing NHS England’s ‘data stewardship public dialogue’.

## Results

Our study included 48,352,770 registered adult patients across 6,389 practices, >98% of total practices in England. A description of patient characteristics of the study population is described in Table 2. We developed a suite of 11 key measures indicative of clinical activity to inform restoration of NHS care in general practice, in collaboration with a clinical advisory group. These key measures include routine blood tests (cholesterol, liver function, thyroid, full blood count, glycated haemoglobin, renal function), reviews for long term conditions (asthma, chronic obstructive pulmonary disorder (COPD), medication review), cardiovascular disease (CVD) risk assessment, and blood pressure monitoring (which may be recorded for routine monitoring or diagnosis of acute conditions). From January 2019 to December 2021 we identified 447 million recorded events across the 11 key measures.

**Table 2.** Cohort description using the latest recorded value for all adult patients who were registered at a general practice at any point between January 2019 and December 2021.

| Characteristic | Category | Number of adult patients (% of total population) |
| --- | --- | --- |
| Total |  | 48,352,770 (100.0) |
| Age | 18-19 | 1,398,430 (2.9) |
|  | 20-29 | 7,685,615 (15.9) |
|  | 30-39 | 8,753,520 (18.1) |
|  | 40-49 | 7,754,940 (16.0) |
|  | 50-59 | 8,025,250 (16.6) |
|  | 60-69 | 6,316,120 (13.1) |
|  | 70-79 | 5,068,760 (10.5) |
|  | 80+ | 3,350,125 (6.9) |
|  | Missing | 20 (<0.1) |
| Sex | M | 24,002,030 (49.6) |
|  | F | 24,350,740 (50.4) |
| Ethnicity | South Asian | 3,148,455 (6.5) |
|  | Black | 1,333,335 (2.8) |
|  | Mixed | 604,600 (1.3) |
|  | Other | 1,039,730 (2.2) |
|  | White | 27,900,210 (57.7) |
|  | Missing | 14,326,440 (29.6) |
| IMD quintile | Most deprived | 9,352,000 (19.3) |
|  | 2 | 10,061,470 (20.8) |
|  | 3 | 9,788,670 (20.2) |
|  | 4 | 9,379,320 (19.4) |
|  | Least deprived | 9,241,205 (19.1) |
|  | Missing | 530,100 (1.1) |
| Region | East | 5,222,485 (10.8) |
|  | Midlands | 8,931,820 (18.5) |
|  | London | 8,499,335 (17.6) |
|  | North East | 5,897,280 (12.2) |
|  | North West | 7,421,095 (15.3) |
|  | South East | 7,671,845 (15.9) |
|  | South West | 4,706,435 (9.7) |
|  | Missing | 2,455 (<0.1) |
| EHR provider | TPP | 28,765,400 (59.5) |
|  | EMIS | 19,587,370 (40.5) |

IMD: index of multiple deprivation, EHR: electronic health record

### Study measures

For each measure, the top five most commonly used individual codes from each codelist and commonly used codes by EHR provider are presented in Supplementary Table 1.

### Trends and variation in measures

Rates of activity for each key measure, the number of events recorded and the number of unique patients in which these events occurred across the entire study period are shown in Table 3. The number of patients included from January 2019 to December 2021 ranged from 1.16 million for the COPD review measure to 27.77 million patients for the blood pressure monitoring measure, representing 2.60 million and 79.30 million coded events respectively. The median practice level rate per 1000 registered patients at baseline ranged from 1.10 in COPD reviews to 65.03 in blood pressure monitoring.

**Table 3:**
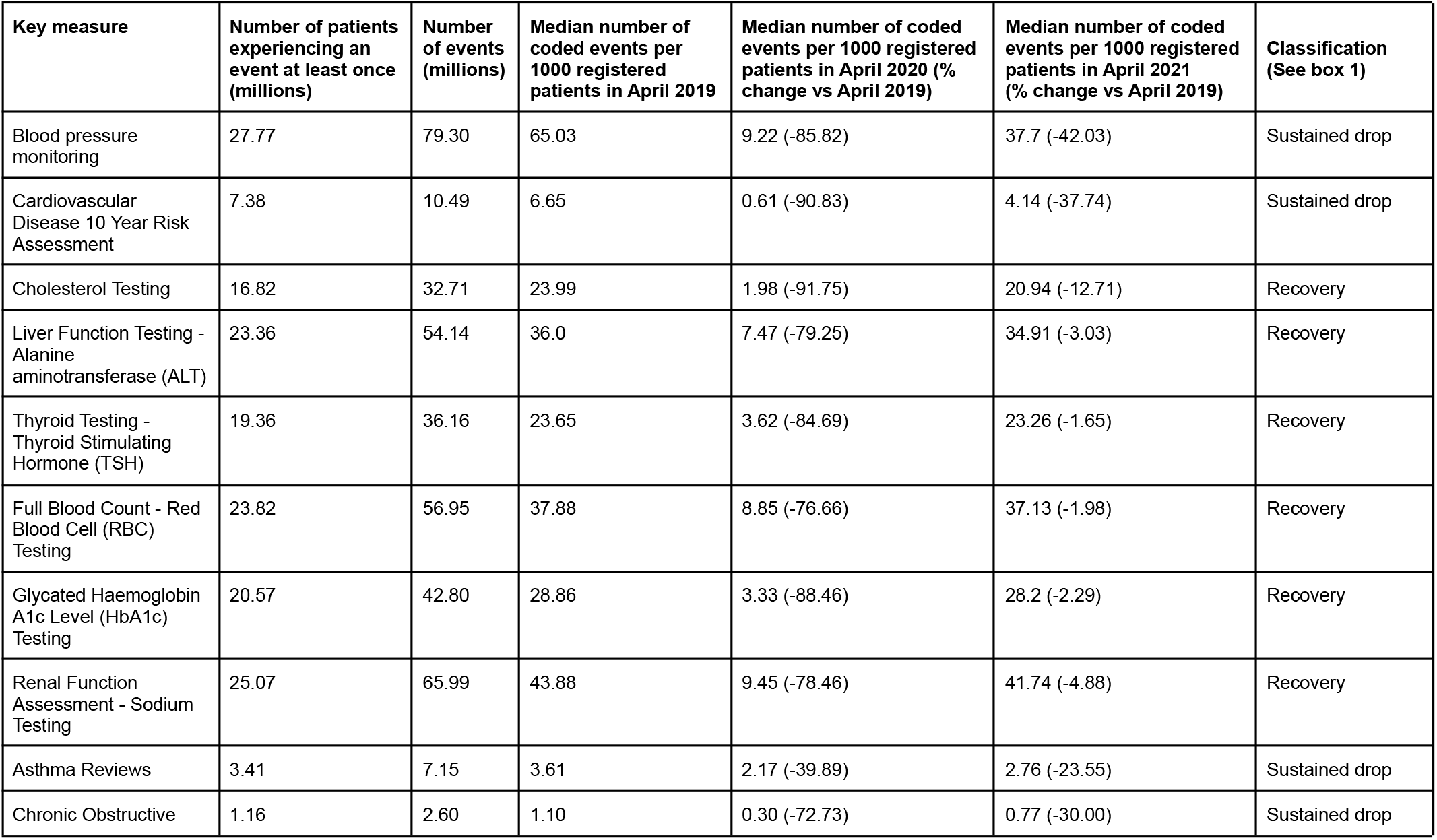
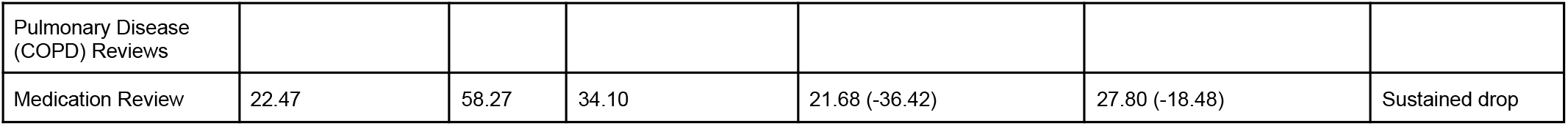
OpenSAFELY NHS SRO Key Measures and their recorded counts and median rate of activity across practices, January 2019-December 2021.

In April 2020, for all measures the median dropped substantially compared to April 2019, ranging from a 91.75% reduction in cholesterol tests (23.99 to 1.98 recorded codes per 1000 registered patients) to a 36.42% reduction in medication reviews (34.10 to 21.68 recorded codes per 1000 registered patients; Table 3). By April 2021 the change in the median compared with April 2019 ranged from a decrease of 42.03% in blood pressure monitoring (April 2019: 65.03, April 2021: 37.70) to a decrease of 1.65% in thyroid testing (April 2019: 23.65, April 2021: 23.26). By April 2021 activity in all six blood monitoring measures had “recovered” to within 15% of baseline, based on the simple SRO classification system. The remaining measures were all classified as having a “sustained drop”. Reviews for asthma and COPD experienced reductions of 39.89% and 72.73% in 2020 respectively. These reductions were sustained in 2021 with rates of 2.76 (−23.55% from baseline) and 0.77 (−30.00% from baseline) in asthma and COPD respectively. Blood pressure monitoring and assessment of cardiovascular 10 year risk were also classified with rates dropping by 42.03% and 37.74% between April 2019 and April 2021.

Figure 1 shows practice level decile charts of the monthly rate per 1000 registered patients for each measure of activity; routinely updating charts are available on the OpenSAFELY Reports website ^18^. Most measures show a similar pattern, a steady rate with wide variation prior to the pandemic with a steep decline in April 2020 during the national lockdown, followed by partial or full recovery over the summer of 2020 and into 2021. Blood pressure monitoring, cardiovascular disease risk assessment and medication reviews continued to show a sustained drop as of December 2021. Blood tests (renal function assessment, cholesterol testing, liver function testing, thyroid function testing, full blood count and glycated haemoglobin) all show a temporary decrease in rates in September 2021. This is likely to be a consequence of a shortage of blood specimen collecting tubes rather than a result of the pandemic, with national guidance to temporarily halt non-clinically urgent blood tests^19^.

**Figure 1.**
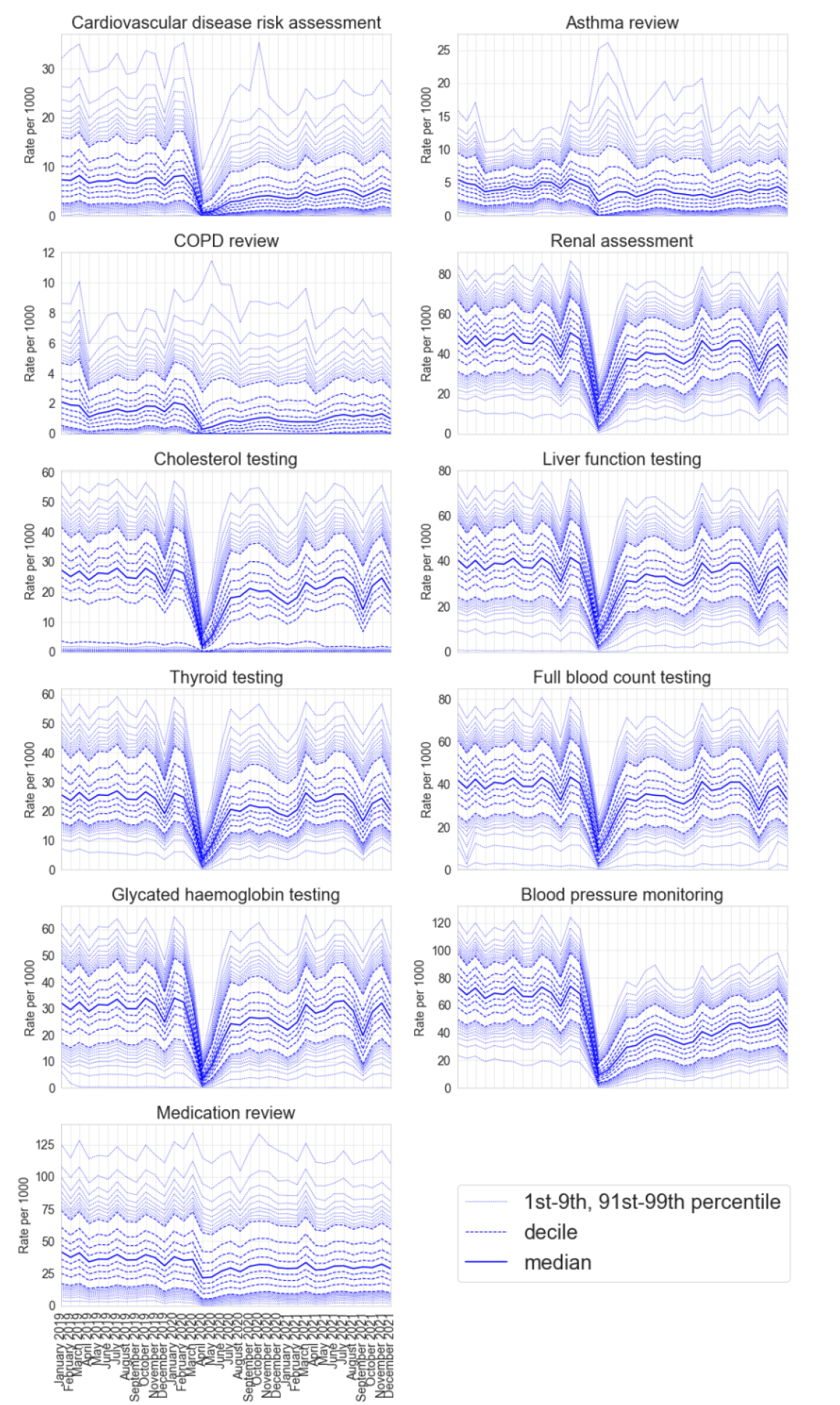
Decile charts of the practice level rate of recorded coding activity per 1000 registered patients in each identified key measure of GP activity between January 2019 and December 2021.

## Discussion

We present 11 key measures of clinical activity and using the OpenSAFELY platform, we executed a federated analysis of changes in these measures throughout the COVID-19 pandemic, across 48 million adults registered at 6,389 general practices in England. These key measures demonstrated substantial changes in clinical activity. Six of the measures recovered to their pre-pandemic baseline within a year of the pandemic, showing a rapid, adaptive response by primary care in the midst of a global health pandemic.

The remaining five measures showed a more sustained drop in activity; asthma and COPD reviews did not recover to their pre-pandemic baseline until around August 2021 and blood pressure monitoring, cardiovascular disease risk assessment and medication reviews had a sustained drop in activity that persisted up to December 2021.

### Strengths and weaknesses

The key strengths of this study are the scale and completeness of the underlying raw EHR data. The OpenSAFELY platform allows federated analysis to be run across the full dataset of all raw, single-event-level clinical events for 57.9 million patients; all patients registered at 99% of all general practices in England. OpenSAFELY can provide data in near-real time, providing unprecedented opportunities for audit and feedback to rapidly identify and resolve concerns around health service activity. We choose when to update the data, and currently update on a weekly basis, meaning the the delay from occurrence of a clinical event to it appearing in the OpenSAFELY platform varies from two to nine days. This is substantially faster than any other source of GP data, including those giving much less complete records.

We also recognise some limitations. With the exception of a small amount of legally restricted data, all occurrences of clinical codes are included, however coded activity may not reflect the true scale and breadth of activity. Codes recorded in general practice do not necessarily indicate unique or new events; for example one patient encounter could generate several similar codes, one patient might have similar diagnoses recorded multiple times over time, or practices might bulk-import information. For each measure, we count no more than one coded event per patient per calendar month, which avoids overcounting where practices use multiple codes to describe a single encounter, but will not account for genuine multiple encounters in a single calendar month. For some of the key measures reflecting routine testing, only test results returned to GPs are included, which will usually exclude tests requested while a person is in hospital and other settings like a private clinic. Our classification system for service change is deliberately simple. A 15% window around the pre-pandemic baseline was chosen as a pragmatic cutoff to highlight these changes. We accept that recovery to the pre-pandemic baseline may not always be expected or appropriate. Finally we are only capturing key measures of clinical activity which does not reflect all clinical care carried out by practices, administrative activity, referral, liaison with other services and other services delivered by general practice.

### Findings in Context

The disruption to health services as a result of the COVID-19 pandemic has been felt globally, with the WHO finding 94% of 135 countries reported some kind of disruption and 48% reported >5% disruption to primary care^20^. Similarly, a systematic review of utilisation of healthcare services during the pandemic reported a 37% reduction in services overall across p20 countries^1^. A study in the UK Clinical Practice Research Datalink (CPRD) of primary care contacts for physical and mental health in the UK showed a considerable drop in activity as a result of national restrictions which only partly recovered by July 2020^3^. Despite changes in evaluated clinical activity, in the winter of 2021 NHS Digital reported that general practice delivered 34.6 million appointments representing a 26% increase (7.1million appointments) in November 2021 compared to pre-pandemic November 2019^21^.

Discussion of the specific causes and reasons for the changes in narrow measures of clinical activity we have described is outside the scope of this paper and is best addressed through quantitative analyses that identify practices in high and low deciles to approach for targeted qualitative interviews with patients and front line staff. However we believe the following broad points may help aid interpretation. Our measures reflect only a few areas of high volume clinical activity; decreases may reflect appropriate prioritisation of other clinical activity as we have found with INR tests^5^ or the delivery of COVID-19 vaccinations^22^. We have previously described how reduced clinical activity can be explained by changes in guidance and financial incentives^6^. For example NHS Health Checks, which are used to detect early signs of high blood pressure, heart disease or type 2 diabetes, were paused during the pandemic; this is likely to explain the sustained drop in activity in cardiovascular disease risk assessment and blood pressure monitoring^23^. However in specific cases this may reflect changes in the style of delivery of a clinical activity, rather than the volume: for example, where patients record their own blood pressure at home since, as we have previously highlighted, home monitoring of blood pressure may not be recorded completely or consistently in GP records. In addition, not all reductions should be interpreted as problematic: as part of the COVID-19 recovery, health systems are aiming to be more resilient, responsive and sustainable^24^; complete recovery may not always be appropriate and reductions in clinical activity across some domains may reflect rational reprioritisation of activity. Where these changes in priority have not been nationally planned, data analyses such as ours may help to rapidly identify the pragmatic changes in prioritisation being made by individual dispersed organisations or people across the healthcare ecosystem before those changes are explicitly surfaced or discussed through other mechanisms.

### Policy Implications and Interpretation

This set of analyses has substantial implications for COVID-19 recovery specifically; the federated analytics platform and framework delivered for these analyses has substantial implications for use of GP data in service improvement and recovery. The COVID-19 pandemic has brought a new challenge for general practice to deliver safe and effective care. Our study, like previous work, has shown substantial changes in clinical activity particularly during the first English lockdown in April 2020 with a quick recovery in certain activities. The measures we have developed with our clinical advisory group are presented here as good measures of clinical activity in general practice and can be easily updated and monitored using our routinely updated dashboards on reports.opensafely.org, although we recommend that they should not be used in isolation as a sole measure of general practice activity. We can expand on these measures to include any measures needed to support NHS England’s ambition to “build back better” as we recover from the COVID-19 pandemic^25,26^. We can update this analysis regularly with extended follow-up time and further measures of activity such as measures defined by the Quality & Outcomes Framework (QOF) and the PINCER medication safety indicators^4,27^ using near-real time data to inform continued progress with NHS service restoration.

More broadly, we have developed an extendable framework for assessing primary care activity and enabling monitoring of service recovery. This framework allows fine-grained analysis over 58 million patient records; analysis that is only possible as we have developed a modular system that allows for federated analytics, where all code written for data curation and analysis is written once, and executed in different locations containing different patients’ data held by different providers. Federated analytics across this scale of NHS EHR data is unprecedented. This approach is efficient: analyses can be easily updated, and expanded, because they are executed in a single framework from re-executable code. It also preserves patient trust: OpenSAFELY was the single most highly trusted COVID-19 data project in a rigorous Citizens Jury sponsored by the NHS and the National Data Guardian^28^. We have also developed interactive “point and click” infrastructure, OpenSAFELY-Interactive (https://interactive.opensafely.org/) to support delivery of dashboards and we are working with NHS England to make this tool available to approved users in order to perform their own similar analyses. OpenSAFELY access is now available to users beyond our own group and we encourage others to use the OpenSAFELY platform and the framework presented here, to develop their own measures of clinical activity. We also plan to develop the functionality for individual practices to receive near real-time feedback on the measures presented here, informing their recovery of service.

### Summary/Conclusion

The COVID-19 pandemic was associated with a substantial change in healthcare activity across the measures we developed. We successfully delivered a secure open source software framework to describe trends and variation in clinical activity across an unprecedented scale of primary care data using federated analytics. We will continue to monitor these changes using our publicly available NHS OpenSAFELY SRO dashboards.

## Data Availability

Access to the underlying identifiable and potentially re-identifiable pseudonymised electronic health record data is tightly governed by various legislative and regulatory frameworks, and restricted by best practice. The data in OpenSAFELY is drawn from General Practice data across England where EMIS and TPP are the data processors.
EMIS and TPP developers initiate an automated process to create pseudonymised records in the core OpenSAFELY database, which are copies of key structured data tables in the identifiable records. These pseudonymised records are linked onto key external data resources that have also been pseudonymised via SHA-512 one-way hashing of NHS numbers using a shared salt. Bennett Institute for Applied Data Science developers and PIs holding contracts with NHS England have access to the OpenSAFELY pseudonymised data tables as needed to develop the OpenSAFELY tools.
These tools in turn enable researchers with OpenSAFELY data access agreements to write and execute code for data management and data analysis without direct access to the underlying raw pseudonymised patient data, and to review the outputs of this code. All code for the full data management pipeline, from raw data to completed results for this analysis and for the OpenSAFELY platform as a whole is available for review at github.com/opensafely-core and github.com/opensafely/sro-measures.

## Administrative

## Acknowledgements

We are very grateful for all the support received from TPP and EMIS throughout this work, and for generous assistance from the information governance and database teams at NHS England and the NHS England Transformation Directorate.

## Conflicts of Interest

All authors have completed the ICMJE uniform disclosure form at www.icmje.org/coi_disclosure.pdf and declare the following: BG has received research funding from the Laura and John Arnold Foundation, the NHS National Institute for Health Research (NIHR), the NIHR School of Primary Care Research, NHS England, the NIHR Oxford Biomedical Research Centre, the Mohn-Westlake Foundation, NIHR Applied Research Collaboration Oxford and Thames Valley, the Wellcome Trust, the Good Thinking Foundation, Health Data Research UK, the Health Foundation, the World Health Organisation, UKRI MRC, Asthma UK, the British Lung Foundation, and the Longitudinal Health and Wellbeing strand of the National Core Studies programme; he is a Non-Executive Director at NHS Digital; he also receives personal income from speaking and writing for lay audiences on the misuse of science. R Conibere declares Honoraria for Work with Primary Care Pharmacy Association. MS is an NIHR funded academic clinical fellow in primary care. MM is NHS GIRFT Senior Clinical Advisor for Pathology.

## Funding

This research used data assets made available as part of the Data and Connectivity National Core Study, led by Health Data Research UK in partnership with the Office for National Statistics and funded by UK Research and Innovation (grant ref MC_PC_20058). In addition, the OpenSAFELY Platform is supported by grants from the Wellcome Trust (222097/Z/20/Z); MRC (MR/V015757/1, MC_PC-20059, MR/W016729/1); NIHR (NIHR135559, COV-LT2-0073), and Health Data Research UK (HDRUK2021.000, 2021.0157).

BG has also received funding from: the Bennett Foundation, the Wellcome Trust, NIHR Oxford Biomedical Research Centre, NIHR Applied Research Collaboration Oxford and Thames Valley, the Mohn-Westlake Foundation; all Bennett Institute staff are supported by BG’s grants on this work. BMK is employed by NHS England and seconded to the Bennett Institute.

The views expressed are those of the authors and not necessarily those of the NIHR, NHS England, UK Health Security Agency (UKHSA) or the Department of Health and Social Care.

Funders had no role in the study design, collection, analysis, and interpretation of data; in the writing of the report; and in the decision to submit the article for publication.

## Information governance and ethical approval

NHS England is the data controller; EMIS and TPP are the data processors; and the key researchers on OpenSAFELY are acting on behalf of NHS England. This implementation of OpenSAFELY is hosted within the EMIS and TPP environments which are accredited to the ISO 27001 information security standard and are NHS IG Toolkit compliant;^29,30^ patient data has been pseudonymised for analysis and linkage using industry standard cryptographic hashing techniques; all pseudonymised datasets transmitted for linkage onto OpenSAFELY are encrypted; access to the platform is via a virtual private network (VPN) connection, restricted to a small group of researchers; the researchers hold contracts with NHS England and only access the platform to initiate database queries and statistical models; all database activity is logged; only aggregate statistical outputs leave the platform environment following best practice for anonymisation of results such as statistical disclosure control for low cell counts.^31^ The OpenSAFELY research platform adheres to the obligations of the UK General Data Protection Regulation (GDPR) and the Data Protection Act 2018. In March 2020, the Secretary of State for Health and Social Care used powers under the UK Health Service (Control of Patient Information) Regulations 2002 (COPI) to require organisations to process confidential patient information for the purposes of protecting public health, providing healthcare services to the public and monitoring and managing the COVID-19 outbreak and incidents of exposure; this sets aside the requirement for patient consent.^32^ Taken together, these provide the legal bases to link patient datasets on the OpenSAFELY platform. GP practices, from which the primary care data are obtained, are required to share relevant health information to support the public health response to the pandemic, and have been informed of the OpenSAFELY analytics platform.

This study was approved by the Health Research Authority (REC reference 20/LO/0651).

## Guarantor

BG is guarantor.

## Contributorship

**Conceptualization**: L.F., H.J.C., R. Croker, C.E.M., B.M., and B.G.

**Data curation**: P.I., C.E.M., A.M., S.B., G.H., C.B., D.E., I.D., T.W., J. Cockburn, S.D., R.M.S., J.P., F.H., S.H., J. Cockburn, S.O.H., A.E., R.J., D.A., P.G., A.F., and N.P.

**Formal analysis**: L.F., H.J.C., R. Croker, and B.M.

**Funding acquisition**: F.H. and B.G.

**Investigation**: L.F., H.J.C., R. Croker, M. Wiedemann, J. Massey, C.E.M., and B.M.

**Methodology**: L.F., H.J.C., R. Croker, L.E.M.H, R.H, C.E.M., A.J.W., O.M., T.L., M. Wood, M.M., M.S., R. Conibere, W.B., H.S., C.D., K.C., W.H., A.G., and B.M.

**Resources**: P.I., C.E.M., A.M., S.B., G.H., C.B., D.E., I.D., T.W., J. Cockburn, S.D., R.M.S., J.P., F.H., S.H., J. Cockburn, S.O.H., A.E., R.J., D.A., P.G., A.F., and N.P.

**Software**: P.I., C.E.M., A.M., S.B., G.H., C.B., D.E., I.D., T.W., J. Cockburn, S.D., R.M.S., S.H., J. Cockburn, S.O.H., A.E., R.J., D.A., P.G., A.F., and N.P.

**Supervision**: B.G.

**Visualisation**: L.F., H.J.C., R. Croker, and B.M.

**Writing - original draft**: L.F., H.J.C., R. Croker, V.S., C.W., A.B., and B.M.

**Writing - review & editing**: L.F., H.J.C., R. Croker, M. Wiedemann, V.S., C.W., A.B., L.E.M.H, R.H, J. Massey, P.I., C.E.M., A.J.W., J. Morley, A.M., S.B., G.H., O.M., T.L., M. Wood, M.M., M.S., R. Conibere, W.B., H.S., C.D., K.C., C.B., D.E., I.D., T.W., J. Cockburn, S.D., R.M.S., W.H., A.G., J.P., F.H., S.H., S.O.H., A.E., R.J., D.A., P.G., A.F., N.P., B.M., and B.G.

## Notes

### Author Declarations

This study was approved by the Health Research Authority (REC reference 20/LO/0651).

### Summary of Updates

Adds in the following authors who were missing: Kiren Collison. Fixes incorrect population counts stated in the manuscript. All references to a population count of 53 million have been replaced with 48 million.

