## Supplementary Material for "Eleven key measures for monitoring general practice clinical activity during COVID-19 using federated analytics on 48 million adults’ primary care records through OpenSAFELY" (from another version of this preprint: version 1, 2022-10-17)

**Supplementary Table 1. Counts of the five most commonly used codes within each measure codelist between January 2019 and April 2021.**

| Measure | Code | Description | Proportion of codes, TPP (%) | Proportion of codes, EMIS (%) | Total Proportion of codes (%) |
|---|---|---|---|---|---|
| Blood pressure monitoring | 72313002 | Systolic arterial pressure | 95.6 | 96.7 | 96.2 |
| | 413606001 | Average home systolic blood pressure | 2.9 | 2.3 | 2.5 |
| | 407554009 | Sitting systolic blood pressure | 0.5 | 0.3 | 0.4 |
| | 400974009 | Standing systolic blood pressure | 0.5 | 0.3 | 0.3 |
| | 314445006 | Average night interval systolic blood pressure | 0.1 | - | 0.1 |
| | 314449000 | Average 24 hour systolic blood pressure | - | 0.2 | - |
| Cardiovascular Disease 10 Year Risk Assessment | 718087004 | QRISK2 cardiovascular disease 10 year risk score | 98.9 | 95.1 | 96.5 |
| | 763244005 | QRISK cardiovascular disease 10 year risk calculator score | 0.8 | 3.7 | 2.6 |
| | 1085871000000105 | QRISK3 cardiovascular disease 10 year risk calculator score | 0.3 | 0.9 | 0.6 |
| | 809311000000105 | Cardiovascular disease risk assessment by third party | <0.1 | 0.3 | 0.2 |
| | 752451000000100 | Framingham coronary heart disease 10 year risk score | - | 0.1 | 0.1 |
| | 752451000000100 | Joint British Societies cardiovascular risk score | <0.1 | - | - |
| Cholesterol Testing | 1005671000000105 | Serum cholesterol level | 99.1 | 96.9 | 97.9 |
| | 1017161000000104 | Plasma total cholesterol level | 0.8 | 3.1 | 2.0 |

| | | | | | |
|---|---|---|---|---|---|
| | 850981000000101 | Cholesterol level | 0.2 | <0.1 | 0.1 |
| | 853681000000104 | Total cholesterol level | - | <0.1 | <0.1 |
| | 365794002 | Finding of serum cholesterol level | - | <0.1 | <0.1 |
| | 395153009 | Pre-treatment serum cholesterol level | <0.1 | - | - |
| Liver Function Testing - Alanine aminotransferase (ALT) | 1018251000000107 | Serum alanine aminotransferase level | 96.4 | 95.6 | 96.0 |
| | 1013211000000103 | Plasma alanine aminotransferase level | 3.5 | 4.2 | 3.8 |
| | 250637003 | Alanine aminotransferase - blood measurement | 0.1 | 0.3 | 0.2 |
| | 34608000 | Alanine aminotransferase measurement | 0.1 | - | <0.1 |
| | 201321000000108 | Serum alanine aminotransferase level | - | <0.1 | <0.1 |
| | 390961000 | Plasma alanine aminotransferase level | - | <0.1 | - |
| Thyroid Testing (TSH) | 1022791000000101 | Serum thyroid stimulating hormone level | 97.6 | 97.1 | 97.3 |
| | 1022801000000102 | Plasma thyroid stimulating hormone level | 2.3 | 2.7 | 2.5 |
| | 1027151000000105 | Thyroid stimulating hormone level | 0.1 | 0.2 | 0.2 |
| | 61167004 | Thyroid stimulating hormone measurement | - | <0.1 | <0.1 |
| | 313440008 | Measurement of serum thyroid stimulating hormone | - | <0.1 | <0.1 |
| Full Blood Count - Red Blood Cell (RBC) Testing | 1022451000000103 | Red blood cell count | 100.0 | >99.9 | >99.9 |
| | 365625004 | Finding of red blood cell count | - | <0.1 | <0.1 |
| | 14089001 | Red blood cell count | - | <0.1 | <0.1 |
| Glycated Haemoglobin A1c | 999791000000106 | Haemoglobin A1c level - IFCC standardised | 98.9 | 94.5 | 96.5 |

| | | | | | |
|---|---|---|---|---|---|
| Level (HbA1c) | 1003671000000109 | Haemoglobin A1c level | 1.1 | 5.5 | 3.5 |
| | 43396009 | Hemoglobin A1c measurement | 0.1 | <0.1 | <0.1 |
| | 365845005 | Hemoglobin A1C - diabetic control finding | <0.1 | <0.1 | <0.1 |
| | 313835008 | Haemoglobin A1c measurement aligned to the Diabetes Control and Complications Trial | - | <0.1 | <0.1 |
| Renal Function Assessment - Sodium Testing | 1000661000000107 | Serum sodium level | 97.8 | 96.8 | 97.2 |
| | 1017381000000106 | Plasma sodium level | 2.2 | 3.2 | 2.8 |
| Asthma Reviews | 394700004 | Asthma annual review | 63.6 | 60.8 | 62.1 |
| | 394720003 | Asthma medication review | 17.4 | 8.4 | 12.5 |
| | 401182001 | Asthma monitoring by nurse | 9.2 | 8.6 | 8.9 |
| | 270442000 | Asthma monitoring check done | - | 11.0 | 5.9 |
| | 401183006 | Asthma monitoring by doctor | - | 5.1 | 2.7 |
| | 394701000 | Asthma follow-up | 4.8 | - | - |
| | 390877003 | Step up change in asthma management plan | 2.6 | - | - |
| Chronic Obstructive Pulmonary Disease (COPD) Reviews | 394703002 | Chronic obstructive pulmonary disease annual review | 93.9 | 98.7 | 96.5 |
| | 760621000000103 | Chronic obstructive pulmonary disease 6 monthly review | 3.5 | 0.9 | 2.1 |
| | 760601000000107 | Chronic obstructive pulmonary disease 3 monthly review | 2.7 | 0.4 | 1.4 |
| Medication Review | 314530002 | Medication review done | 69.3 | 8.0 | 34.7 |
| | 182836005 | Review of medication | 3.0 | 42.7 | 25.4 |
| | 88551000000109 | Medication review with patient | 2.9 | 19.6 | 12.3 |

| | | | | | |
|---|---|---|---|---|---|
| | 93311000000106 | Medication review of medical notes | - | 12.5 | 7.1 |
| | 394720003 | Asthma medication review | 2.5 | 2.0 | 2.3 |
| | 719329004 | Medication review done by pharmacist | 3.1 | - | - |

“-”: code outside the top five codes
